# Prioritizing the first doses of SARS-CoV-2 vaccine to save the elderly: the case study of Italy

**DOI:** 10.1101/2021.02.26.21252538

**Authors:** Giuseppe Pontrelli, Giulio Cimini, Marco Roversi, Andrea Gabrielli, Gaetano Salina, Stefania Bernardi, Francesca Rocchi, Alessandra Simonetti, Carlo Giaquinto, Paolo Rossi, Francesco Sylos Labini

## Abstract

Many countries are currently facing high mortality caused by the circulation of SARS-CoV-2 among the elderly not yet vaccinated. Vaccine shortage poses relevant challenges to health authorities, called to act in a timely manner, and with scarcity of vaccine, and data. We have developed a model for estimating the impact of vaccination on the mortality of the elderly following a schedule of mRNA SARS-CoV-2 vaccine that prioritize first dose administration, as alternative to the standard schedule of two doses administered 3 to 4 weeks apart. We studied the Italian scenario, considering it representative of other Countries facing similar conditions in terms of virus circulation, mortality, and vaccine shortage, in the period from February, 10 to April, 14 2021. Under different conditions of quantity of vaccine administration, the schedule prioritizing first doses showed always significant increase of protected individuals, and a decrease of deaths, up to 19.8% less than the standard schedule. These findings support the vaccination option of prioritizing first dose in the elderly until vaccine supplies are adequate.

## Main document

Despite the restrictive measures adopted worldwide, the daily count of infections and deaths from COVID-19, remains high and unbearable. In western countries the highest death toll has been paid by the elderly: in the case of Italy, the latest bulletin of the Italian National Institute of Health (Istituto Superiore di Sanità, ISS), updated on February 17, 2021, reported that 62.1% of the 93,074 deaths due to the pandemic were over 80 years old. The vaccination campaign has therefore prioritized this age group, whose immunization in Italy began on Monday, February 8, with some difficulties due to various delays in the supply of the two approved mRNA vaccines (Pfizer/BNT Biotech and Moderna) dedicated to this cohort. The Italian Strategic Plan for anti-SARS-CoV-2/COVID-19 Vaccination has been consequently adjusted several times^1^.

The recommendation derived from registrative trials is that administration of the mRNA vaccines should be in two doses, spaced three or four weeks apart: the efficacy of preventing symptomatic COVID-19 in clinical trials was of 94.8% and 94.1% for the two vaccine, Pfizer/BNT Biotech and Moderna respectively^2,3^. However, when excluding cases of infection in the first 14 days after the first dose (the time needed for the activation of an effective immune response against the vaccine antigen), the same trial studies have shown a good effectiveness of the first dose alone: 92.6%^4^ and 92.1%^3^ for Pfizer/BNT Biotech and Moderna vaccine, respectively. A recent Israeli study also estimated a first dose efficacy of 85% (95%CI 71-92) in reducing symptomatic COVID-19 cases^5^. This data was recently confirmed in a study in Scotland on Healthcare Workers^6^.

All these data refer to short-term efficacy, but show efficacy above the threshold considered by the EMA (European Medicines Agency) guidelines. The available data do not demonstrate that the vaccination prevents transmission, yet we know that the vaccine reduces the symptomatic forms of COVID-19, and as a result a decrease in both the number of severely affected patients requiring admission to the ICU and deaths is expected.

Based on epidemiological data and the scarcity of vaccine doses compared to the amount needed to protect the population, the United Kingdom has adopted the strategy of postponing the administration of the second dose to 12 weeks after the first^7^. The goal is self-evident: to protect as many people as possible as soon as possible, while waiting for a better supply of vaccines.

In Italy the available doses of the two mRNA vaccines are much lower than those needed to immunize the entire population or even the over-80s in a short time. This situation is putting Italy, and other countries like the U.S., in front of a question similar to that faced by the United Kingdom: if the first objective remains that of saving the greatest number of lives, why not delay the second doses until all high-risk subjects have been vaccinated with at least one dose?

Stanley Plotkin, asked this question in an article that appeared recently^8^, and answers the question favorably, considering not only the apparent correlation between protection and low antibody levels after a single-dose administration of mRNA vaccine, as demonstrated in some studies, but also the relative efficacy of other vaccines, especially anti-hepatitis B, when administered at prolonged intervals^2,9,10^. Finally, he reminds us how memory B cells develop properly following administration of mRNA vaccines, supporting the idea that further enhancement of antibody production is stimulated by a second dose of vaccine given even six months after the first^11^. Along the same lines, recent modelling studies on SARS-CoV-2 showed the effectiveness of one dose vaccination in containing the pandemic more rapidly^12^.

Here we provide a computation of the expected benefits to support the choice of the best vaccination strategy. We focused our analysis on the Italian cohort of 4,442,048 people over 80 years old, who should be vaccinated up to March 31, 2021, according to the current National Vaccination Plan^1^. We formulated a simple effective model that estimates the number of protected individuals and deaths under different scenarios assuming a rate of weekly vaccine administration. Let us consider, for example, a scenario with 800,000 doses (about 18% of the cohort) of mRNA vaccine administered each week.

Under the standard schedule of two doses 3 and 4 weeks apart, a part of the doses will be reserved and used for the second vaccination of subjects already vaccinated, to the detriment of the vaccination of over 80 years old not yet vaccinated, who will have to wait while remaining at risk. Considering the latency of two weeks necessary to evoke an efficient immune response, with this strategy a total of 2.929,000 people will be protected as of April 14, 2021. If we assume a constant weekly mortality rate of 40 fatalities per 100,000 people in this age group (update of February 10, 2021^13^) there will be 7,061 deaths in the cohort by April 14. In contrast, following the alternative schedule, i.e., the strategy of prioritizing first doses before moving on to second doses, in the same time interval 3,727,331 people will be protected and a total of 5,664 deaths in the cohort are expected. That is, a 19.8% reduction of deaths, which notably is independent of the assumed mortality rate. The benefit is evident even when considering a different number of weekly vaccinations (see Table 1 and Figure 2), the simple reason being the number of protected growing at a very different pace in periods when the first doses or the second doses are administered. It should be noted that the strategy of temporarily postponing the second dose could also be applied to other cohorts identified by the vaccination plan as priorities, for example extremely vulnerable individuals, as patients with chronic conditions that pose additional risk of death if infected, to obtain additional benefits, comprehending patients in pediatric age as Pfizer vaccine is authorized over 16 years.

**Table 1.** Comparison between standard and alternative schedule.

|  | **Standard Schedule:**<br>**two doses 3-4 weeks apart.** | **Alternative Schedule:**<br>**priority to first dose.** |
| --- | --- | --- |
| **Doses per week** | 400,000 (approximately 9.0% of cohort) |  |
| Protected at 14/04 | 1,460,000 | 2,240,000 (+53.4%) |
| Deaths 24/2 - 14/4 | 9,750 | 8,854 (-9.2%) |
| Time between doses | 3-4 weeks | 11 weeks |
| **Doses per week** | 600,000 (approximately 13.5% of cohort) |  |
| Protected at 14/04 | 2,190,000 | 3,360,000 (+53.4%) |
| Deaths 24/2 - 14/4 | 8,406 | 7,062 (-16%) |
| Time between doses | 3-4 weeks | 7.5 weeks |
| **Doses per week** | 800,000 (approximately 18.0% of cohort) |  |
| Protected at 14/04 | 2,920,000 | 3,727,33 (+27.6%) |
| Deaths 24/2 - 14/4 | 7,061 | 5,664 (-19.8%) |
| Time between doses | 3-4 weeks | 5.5 weeks |

**Figure 1.**
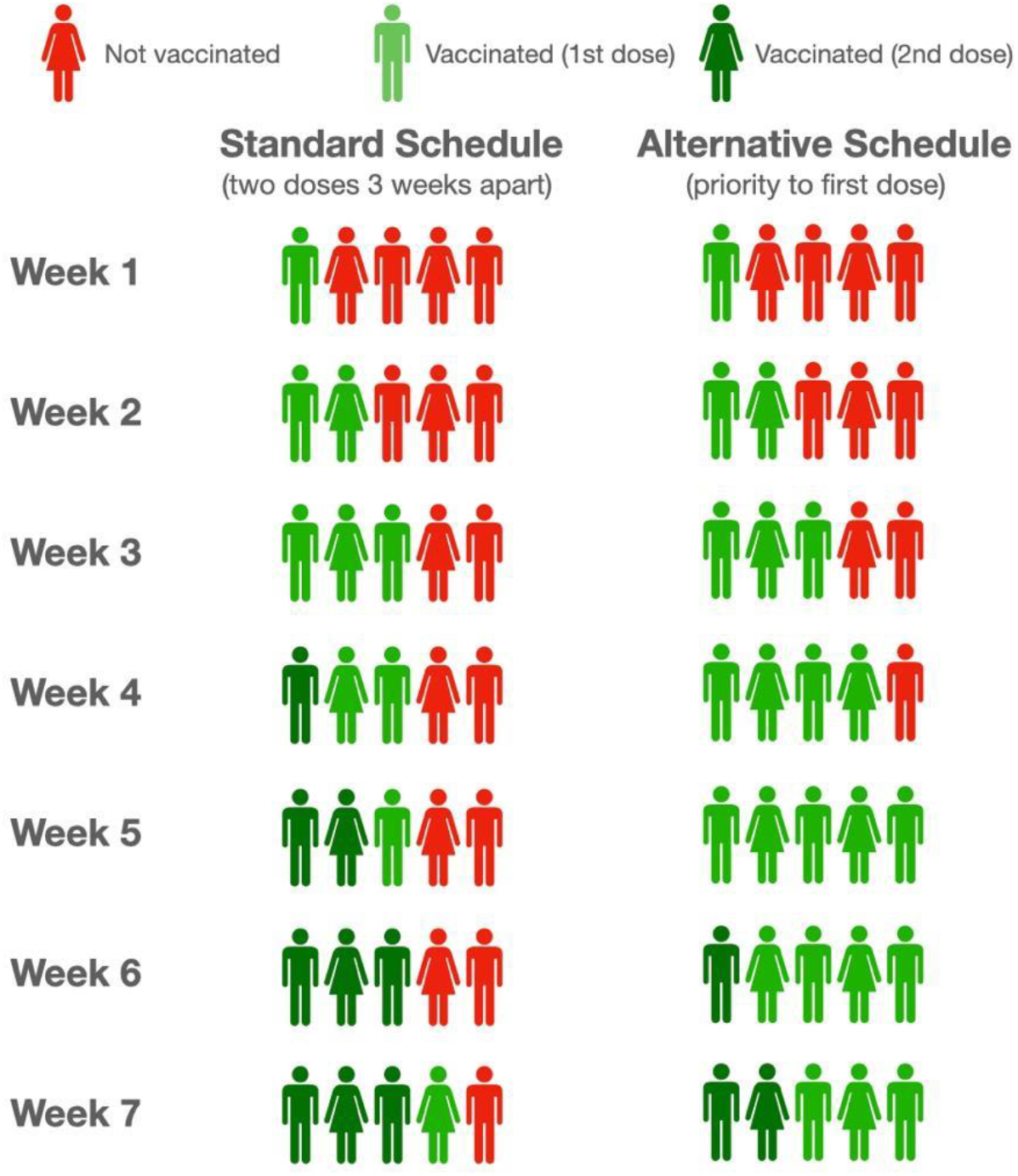
Schematic illustration of standard and alternative schedule of vaccination.

**Figure 2.**
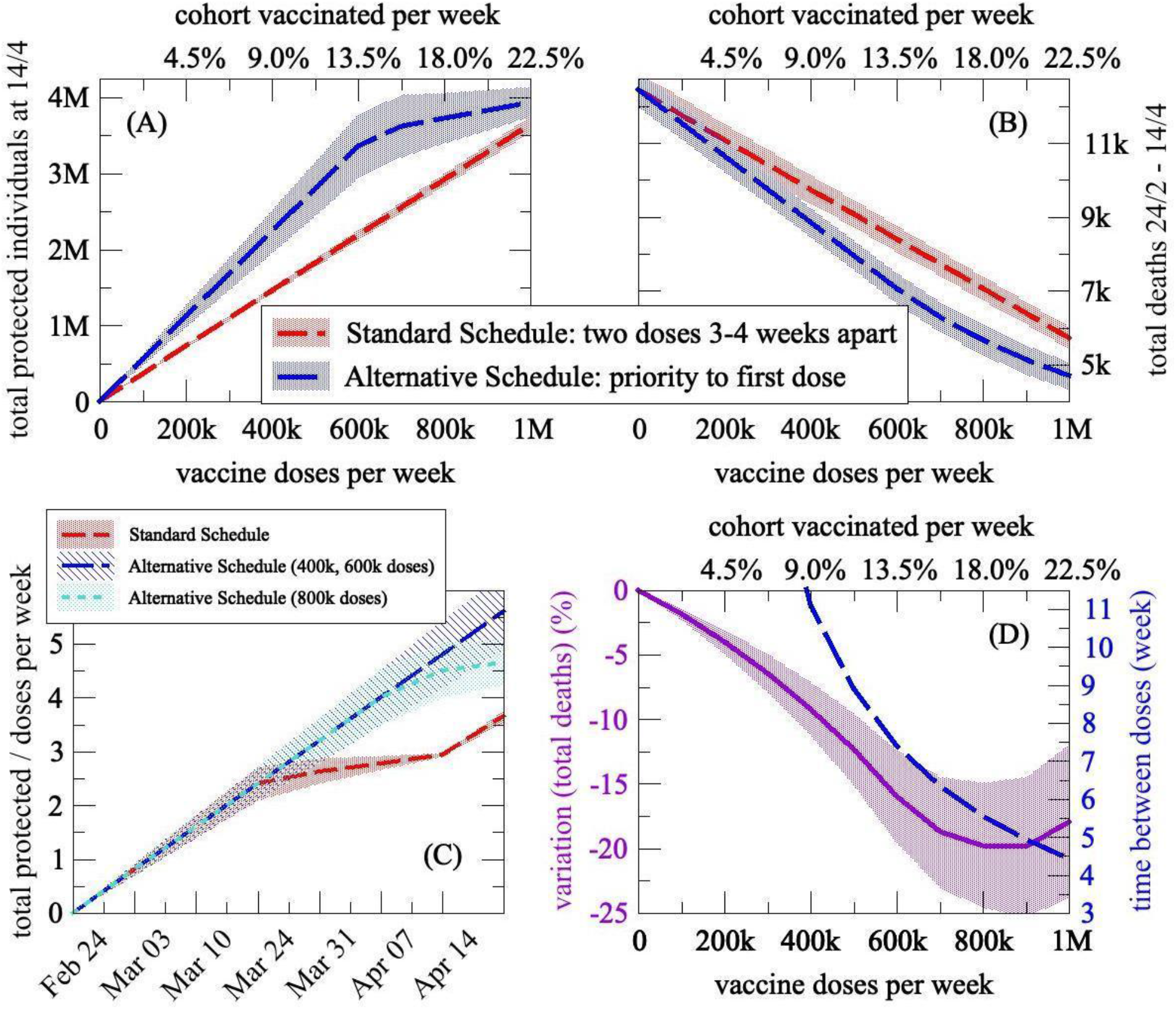
Total protected individuals at April, 14 (A) and total deaths in the period February, 24 - April,14 (B) in the cohort according to the two schedules, as a function of the number of doses administered per week (or equally expressed as the percentage of the cohort vaccinated each week). The alternative schedule allows protection of many more individuals, and converges to the standard one only for an extremely high and improbable, given the number of doses available, number of weekly administrations (about 1 million, see also plot (D)). However, the advantage of the alternative schedule considering deaths is quite large at any time, since it is derived by the advantage in protection accumulated during the time span of the model, see plot (C). (C) Comparison of the standard and alternative schedules (with a different number of weekly doses administered) on *efficiency index*, that is the number of protected individuals for doses administered over time. Protection increases much more slowly in the standard schedule at time of second doses administered. (D) Analysis of the alternative schedule in terms of decrease in deaths as compared with the standard schedule, and effective week in which administration of second doses begins, after the entire cohort has received the first dose of vaccine. In all plots the shaded region denotes the confidence interval derived from the uncertainty associated with the efficacy of first dose and mortality rate.

The U.S. CDC addressed the problem this way stating “There is no maximum interval between the first and second dose for either vaccine (Pfizer/BNT Biotech and Moderna). Therefore, if the second dose is administered >3 weeks after the first Pfizer-BioNTech vaccine dose or >1 month after the first Moderna vaccine dose, there is no need to restart the series”^14^.

It may be argued that a delayed second dose facilitates the emergence of vaccine-resistant variants of the virus. Nonetheless, data available show that vaccines appear protective on variants now circulating in Europe^6,15,16^, and the risk of this emergence is counterbalanced by the advantages of reducing viral circulation, by making more people nonsusceptible to the virus in a shorter amount of time^17^. In any case, in the alternative schedule the interval between the first and second doses is limited if the number of weekly vaccines administered is high (less than 7.5 weeks if more than 600,000).

Based on these findings, we suggest to take in consideration the vaccination option to prioritize first dose in the elderly until vaccine supplies are adequate.

## Model Assumptions

We consider a 7-week period, from February 10 to March 31, counting protected individuals and the deaths from February 24 to April 14 (considering the 2-week period required for coverage activation). Protected individuals in a given week are obtained as the total individuals vaccinated at 1 or 2 doses two weeks earlier, each modulated for the effective coverage provided by the vaccine. Deaths at a given week are then obtained through the weekly mortality parameter applied to the susceptible population (the cohort) after subtracting the protected individuals.

### Model Parameters

- Efficacy of 2nd dose after 14 days: 0.95^2,3^
- Efficacy of 1st dose after 14 days: 0.80±0.10^5^
- Cohort over 80 years of age: 4,442,048
- Available mRNA vaccines on March 31: 8,179,748 (i.e. the share of the Vaccination Plan minus the administrations already performed as of February 10)^1^
- Ratio of available Pfizer/BNT Biotech to Moderna vaccines: 6.77^1^
- Weekly mortality rate (number of deaths on susceptibles) in over 80s in the week 3-10 February 2021 in Italy: 0.00040±0.00004^13^

(*) For Italy, using the full supply of vaccines for the first quarter of 2021 in the 7 weeks of the model would result in slightly over 1 million doses administered per week.

## Data Availability

Original data needed for building the dataset and the model are publicly available, at the links reported in Bibliography.

## Authors’ contributions

Conceptualized and designed the article: GP, GC, MR, AG, GS, FSL. Collected data: GP. Analyzed data: GP, GC. Verified data: GP, GC, MR, AG, GS, FSL. Created the tables and graphs: GC. Drafted and wrote the manuscript: GP, GC, MR. Reviewed the manuscript for important intellectual content: SB, FR, AS, CG, PR. Revised the final manuscript: GP, GC, MR, AG, GS, SB, FR, AS, CG, PR FSL

